# Prepayment meters strongly associated with economic and health deprivation: an observational, cross-sectional study

**DOI:** 10.1101/2022.12.19.22283693

**Authors:** Xuejie Ding, Evelina Akimova, Bo Zhao, Kasimir Dederichs, Melinda C. Mills

## Abstract

**Objectives:** To examine to what extent pre-payment meters (PPMs) are associated with multiple measures of structural economic and health deprivation.

**Design:** Cross-sectional, observational.

**Setting:** England and Wales.

**Data source:** Experimental Lower Layer Super Output Area (LSOA) prepayment electricity meter consumption 2017 is used to map the percentage of PPMs for each LSOA. Number of domestic prepayment meters and meters for 33,332 LSOA regions across England and Wales are linked with multiple sub-national LSOA deprivation and Middle Layer Super Output Area (MSOA) health measures. Smart meters operating in prepayment mode are not included.

**Outcome measures:** Prevalence of PPMs for electricity bills per LSOA, associated with multiple national deprivation measures. In England: fuel poverty; income deprivation children and older people; living environment; barriers to housing and services; crime; health and disability; education, skills and training; employment; income; overall Index of Multiple Deprivation (IMD). In Wales: physical environment; community safety; housing; access to services; education; health; employment; income; overall Welsh Index of Multiple Deprivation (WIMD). Additional outcomes of Tenure Type of Housing (rented social, private; owned) and emergency hospital admissions for chronic obstructive pulmonary disease (England) and all respiratory diseases (Wales).

**Results:** PPM prevalence is strongly correlated (0.62 to 0.63) with fuel poverty, with similar patterns in England and Wales. In England, PPM prevalence was strongly associated with virtually all deprivation indicators, including: lower income (−0.83 [−0.83 to −0.82]), receipt of employment-related benefits (−0.76 [−0.77 to −0.76]), educational disadvantage (−0.70 [−0.70 to −0.69]) and higher levels of health deprivation (−0.65 [−0.65 to −0.63]). Higher PPM prevalence is concentrated in low-income families with children and older individuals, strongly associated with income deprivation affecting children (−0.8 [−0.81 to −0.80]) and older people (−0.79 [−0.80 to −0.79]), social rent (0.67 [0.66 to 0.68]), hospital admissions for respiratory diseases (England 0.69 [0.68 to 0.70]; Wales 0.64 [0.58 to 0.70]), indicating a heightened risk of illness.

**Conclusions:** Instead of blanket policies, interventions on households with PPMs offers an immediate solution to avoid a health crisis and shield vulnerable households from a ‘heat or eat’ dilemma and further exacerbating existing inequalities.

**Strengths and limitations of the study:** This is the first analysis linking structural economic and health inequality to pre-payment meters (PPM) in the current energy crisis.

PPMs are a similar to standard measures of fuel poverty, but a more straightforward proxy for a rapid targeted intervention to avoid a surge in hospital emergency admissions this winter as the most vulnerable households face a ‘heat or eat’ dilemma.

PPMs are concentrated in vulnerable households (children, elderly, out of employment) and highly correlated with socioeconomic deprivation and emergency hospital admissions for respiratory diseases.

PPMs do not include smart meters operating in prepayment mode, underrepresenting the observed level of poverty.

The study includes only England and Wales. Care should be taken when generalising to other areas. We call for the release of PPM data in Scotland and Northern Ireland, to include payment methods in social surveys to evaluate risk for focused interventions.

## Introduction

Global energy prices have soared since mid-2021 due to post-pandemic energy requirements and the Russian invasion of Ukraine that reduced supply to the European energy market. Within seven months of the Russian invasion, European energy wholesale prices skyrocketed by 109% (gas) and 138% (electricity). As the war rages, the energy market remains restricted.[1] The UK energy crisis is estimated to thrust 11 million UK households into fuel poverty within the next year, [2] prompting a ‘humanitarian crisis’ as millions of children fall into fuel poverty.[3] Given that fuel poverty is inherently linked to poor health, this energy crisis is about to turn into a major health crisis.[4] A lesson from the COVID-19 pandemic is that emergency shocks do not impact individuals equally.[5] Rather, structural shocks like the energy crisis have an unequal impact on individuals, exacerbating existing inequalities. A relatively understudied structural inequality in the UK is energy inequality. In 2020, a report showed that lower income households pay in relative terms, £60 *more* per year on energy bills compared to wealthy households.[6] By the end of 2022, the poorest 10% of families could pay 47% of their disposable income on energy costs, while the top 10% households pays 20%.[7] This has fuelled the urgent claims that the poorest households in the UK now need to choose between ‘heat or eat’.

The widening gap in energy inequality in the UK is largely attributed to the structure of payment methods. The most common and effective manner of paying energy bills is via direct debit. There is also the ‘pay as you go’ method, where people use prepayment meters (PPMs) to pay for what they use in advance. PPMs have often been framed positively in that they afford people the control to ‘choose’ their payment methods.[8] In the majority of cases, however, the poorest and most indebted households are forced to opt into PPM because their landlord or energy suppliers have imposed PPMs to avoid debt. Around 4.3 million in the UK currently use a PPM for their energy bills.[9] Recent reports show that around an additional half million could be forced to change from direct debit payment to PPMs by the end of 2022, due to a growing parallel debt crisis.[10] Households on PPMs are usually the most disadvantaged, vulnerable, and have less disposable income for other essentials. In fact, 33% of low-income households in the UK use a PPM or pay in receipt of a bill, whereas 20% of all households in the UK use these pre-payment methods.[11] PPMs also exacerbate poverty since prepayment tariffs are in general more expensive compared to direct debit consumers, which result in paying a ‘poverty premium’. Without effective and urgent policy interventions, PPMs, the energy crisis, and cold weather this winter will push a large group of already vulnerable low-income households into a ‘heat or eat’ dilemma and further exacerbate social inequalities.[12]

Previous research has focused on fuel poverty, measured by the Low Income Low Energy Efficiency (LILEE) indicator.[13] A household is considered fuel poor if: 1) they are living in a property with a fuel poverty energy efficiency rating of band D or below; 2) when after they spend the required amount to heat their home, they are left with a residual income below the official poverty line. While this indicator accounts for household income, household energy requirements and fuel prices to determine whether a household is fuel poor, we contend that PPMs are a more straightforward and efficient indicator to immediately target the fuel poor. We provide evidence that PPMs are tightly associated with the definition of fuel poverty as well as a variety of deprivation domains, including health indicators, at fine-grained local and regional levels.

### Background of prepayment meters

Compared to other European countries, PPMs are widely used in the UK. The number of consumers using PPMs doubled from 7% in 1996 to 15% in 2019.[14,15] Prepayment meters (PPM) are a type of energy payment method that requires the customer to pay for their energy use in advance. Within Gas and Electricity Markets, PPM customers are more vulnerable than direct debit customers.[16] First, PPM users are more likely to have pre-existing debts.[17] PPMs are usually installed into households that hold a debt with energy suppliers, with the aim to ‘help’ households to more effectively manage debt and budgets. Some landlords also insist on PPMs being installed in rental properties to minimise the risk of tenants running into energy debt. Second, PPM users pay more per kilowatt/hour since the best energy deals are unavailable to them. OFGEM[18] published details of average standing charges, showing that from 01 October 2022, direct debit customers pay 46p/day to their electricity supplier and 28p for gas while for PPM customers typically pay 50p/day for electricity and 37p for gas (depending on the supplier and area of residence). For cash and cheque prepayment customers, standing charges would be 51p for electricity and 32p for gas per day[19] in additional to any transactional costs (e.g., time and travel to outlets to purchase credit).[20]

From October 2022, as part of the Energy Bills Support Scheme to deal with the energy crisis, the UK government provided a £400 non-repayable discount on energy bills over 6 months until end March 2023 for all households across Great Britain. The discount works smoothly for households paying energy bills via standard credit, payment card and direct debit, as they receive an automatic deduction to their bills. For traditional PPM customers, Energy Bill discount vouchers from the first week of each month are provided, via SMS text, email or post. The support scheme is thus the least friendly to the most vulnerable customers, since they need to actively redeem the Energy Bill discount voucher at their usual top-up point or Post Office branch. If there is existing debt on the PPM, part of the discount would first be applied to that.

PPM customers are more likely to become or remain trapped in fuel poverty, which has deep consequences.[21] Supplier disconnection or customer self-disconnect from the energy supply because of failure to recharge the PPM in time interrupts power usage, resulting in cold homes and fuel poverty. Although empirical evidence of PPMs and health and social outcomes is scarce, the adverse consequence of living with fuel poverty has been widely noted. Fuel poverty and living in cold temperatures is associated with physical health risks, poor mental health and social outcomes.[22] Burlinson and colleagues found lower fruit and vegetable consumption among PPM customers, suggesting they face a ‘heat or eat’ dilemma.[12] Exposure to cold indoor temperatures and early exposure to respiratory illness have also been shown to have an adverse impact on young children, with lingering impacts over their lifetime.[22] Cold indoor temperatures are also associated with cardiovascular diseases and mental health deterioration, along with emergency hospitalisations.[23] Cold indoor temperatures were also linked to high blood pressure, a key risk factor linked to multiple detrimental health outcomes.[24] Fuel poverty has a profound impact on people with disabilities and serious illness, since the equipment that supports the daily life of the disabled and those will chronic and serious care needs often requires more energy than an average household. Individuals with chronic diseases are prone to feeling cold, placing them at an even higher health risk if they lack heat.

Our research is the first to provide empirical evidence to demonstrate that PPMs are associated with fuel poverty and a variety of socioeconomic and health deprivation indicators. Exploring PPMs can be a means of investigating the outcomes of fuel poverty among a specific and easily identifiable group, who require urgent support to survive the winter.

### Data, Measures and Methods

#### Data and Methods

We first used Experimental Lower Layer Super Output Area (LSOA) Prepayment Electricity Meter Consumption 2017 data to map the percentage of PPMs for each LSOA.[25] 1,393 of 32,844 (4%) LSOA in England and 28 of 1909 (1%) LSOA in Wales have missing values in PPM. To explore the missingness, we plot the distribution of IMD for LSOA with missing values in PPM in England and Wales (online supplemental Figure S1) and found that missingness is concentrated in the least deprived areas due to disclosure control – LSOAs are coded a missing PPM value if they contain less than 6 meters. It is worth noting that the data does not include smart meters operating in prepayment mode, suggesting that results may underestimate fuel poverty. Our estimation of sub-national percentage of prepayment electricity meters only contains meters that have consumption between 100 kWh and 100,000 kWh and have a domestic meter profile.[25] We used the following equation to calculate the proportion of prepayment (PPM) electricity meters per LSOA: % PPM per LSOA:

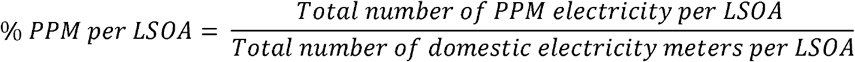

We then link the PPM data with multiple sub-national deprivation and health indices[26,27] to analyse the association between PPMs and geographical inequalities in England and Wales using Pearson correlation:

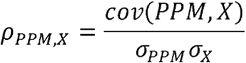

where X is a vector of deprivation and health indices. *ρ*_*PPM,X*_ is the Pearson correlation coefficient for PPM and X. *cov*(*PPM, X*) is the covariance of variables PPM and X. *σ*_*PPM*_ is the standard deviation of PPM. *σ*_*X*_ is the standard deviation of X.

Housing tenure type, proportion of multi-glazing and house year data (reported in online supplement Table S1) were obtained from Energy Performance Certificate.[28] The EPC database was at the postcode level, and we linked it with all local authorities at the LSOA level in England and Wales.[29] The data preparation procedure included removing outliers and duplicates. The cleaned dataset contains the number of properties for each EPC rating per LSOA, number of properties for each tenure type per LSOA, number of properties at construction period per LSOA and proportion of multi-glazed windows per LSOA. For frequencies less than ten, we rounded up figures. The dataset is available in the supplementary materials for public use (online supplementary Figure S2).

We first calculated the proportion of prepayment (PPM) electricity meters per LSOA. This is then associated with multiple national deprivation and health indices to analyse the association (Pearson correlation) between PPMs and geographical inequalities in England and Wales. We measure multiple types of deprivation for small areas (LSOA) and identify the areas with the highest concentrations of the particular type of deprivation. When an index is applied, the small areas (LSOAs) are thus ranked from the most to least deprived, allowing us to map and examine geographic inequalities in England and Wales. In a second analysis, we compare PPM prevalence per LSOA by housing tenure type (rented social housing, rented private housing, owner-occupied). In a third analysis we examine PPM prevalence per Middle Layer Super Output Area (MSOA) by emergency admissions for Chronic Obstructive Pulmonary Disease (COPD) (England) and for all respiratory diseases (Wales). In addition, we examine whether PPM is associated housing quality.

### Measures

Due to different systems, the measures of deprivation differ slightly between England and Wales. For England, we use the English Indices of Deprivation (ID), which provides rankings a means of identifying the most and least deprived areas (LSOAs) and to compare in relative terms whether one area is more deprived than another. Higher ranked areas are the least deprived areas, whereas the lower ranked areas are the most deprived.

The deprivation indices for England are measured by the following indicators. *Fuel poor* is the Low Income Low Energy Efficiency (LILEE) indicator. A household is fuel poor if: 1) they are living in a property with a fuel poverty energy efficiency rating of band D or below; 2) when they spend the required amount to heat their home, they are left with a residual income below the official poverty line. *IDAOPI (Income Deprivation Affecting Older People Index)* measures the proportion of all those aged 60 or over who experience income deprivation. It is a subset of the Income Deprivation Domain that measures the proportion of the population in an area experiencing deprivation relating to low income. *IDACI (Income Deprivation Affecting Children Index)* measures the proportion of all children aged 0 to 15 living in income deprived families. It is a subset of the Income Deprivation Domain which measures the proportion of the population in an area experiencing deprivation relating to low income.

*Living Environment deprivation* measures the quality of the local environment. The indicators fall into two sub-domains. The ‘indoors’ living environment measures the quality of housing and the ‘outdoors’ living environment contains measures of air quality and road traffic accidents. *Barriers to Housing and services deprivation* measures the physical and financial accessibility of housing and local services including the physical proximity of local services and issues relating to access to housing such as affordability. *Crime deprivation* measures the risk of personal and material victimisation at local level. *Health deprivation and disability* measures the risk of premature death and the impairment of quality of life through poor physical or mental health. *Education, skills and training deprivation* measures the lack of attainment and skills in the local population. *Employment deprivation* measures the proportion of the working-age population involuntarily excluded from the labour market. This includes people who would like to work but are unable to do so due to unemployment, sickness or disability, or caring responsibilities. *Income deprivation* measures the proportion of people within small areas or neighbourhoods in England who are income deprived. *Index of Multiple Deprivation (IMD*) classifies the relative deprivation of small areas at the LSOA level. Multiple components of deprivation are weighted with different strengths and compiled into a single score of deprivation. These include and are weighted (shown in brackets) by: income (22.5%), employment (22.5%), education (13.5%), health (13.5%), crime (9.5%), barriers to housing and services (9.3%) and living environment (9.3%).[26]

In Wales, we drew from the Welsh measures of multiple deprivation for small areas,[30] which identifies areas with the highest concentrations of different types of deprivation. *Physical environment deprivation* measures factors in the local area that may impact the wellbeing or quality of life, consisting of three weighted domains of: air quality, flood risk and green space. *Community safety deprivation* measures deprivation relating to living in a safe community and actual experience of crime and fire in addition to perceptions of safety in the local area. It includes six weighted domains of: fire incidents and police recorded criminal damage, violent crime, anti-social behaviour, burglary and theft. *Housing deprivation* measures inadequate housing, in terms of physical and living conditions and availability. *Living condition* refers to the suitability of the housing for its inhabitant(s), such as health and safety and necessary adaptations. It consists of two equally weighted indicators of overcrowding measures the percentage of people living in overcrowded households and poor housing quality, measured by the likelihood of housing being in disrepair or containing serious hazards (for example, risk of falls or cold housing). *Access to services deprivation* measures deprivation as a result of a household’s inability to access a range of physical and online services online considered necessary for day-to-day living. This covers both material deprivation (e.g., not being able to get food) and social aspects of deprivation (e.g., not being able to attend afterschool activities). Poor access to services is a factor that compounds other types of deprivation that exist within an area.

*Education deprivation* captures the extent of deprivation relating to education, training and skills and reflects educational disadvantage within an area in terms of lack of qualifications or skills. *Health deprivation* is a measure of broader range health conditions (age-sex standardised) of people with a GP-recorded diagnosis of chronic conditions. It includes seven weighted indicators of limited long-term illness (e.g., coronary heart disease, Chronic Obstructive Pulmonary Disease), premature deaths (under age 75), GP-recorded mental health conditions (e.g., depression, anxiety, dementia), cancer incidence, low birth weight and children aged 4-5 who are obese. *Employment deprivation* measures the percentage of working-age population in employment deprivation (in receipt of employment-related benefits). It therefore includes involuntary exclusion of the working-age population from work, including those who cannot work due to ill-health or who are unemployed, but actively seeking work. *Income deprivation* measures the percentage of people in income deprivation (in receipt of income-related benefits and tax credits). *The Welsh Index of Multiple Deprivation (WIMD)* is the Welsh Government’s official measure of relative deprivation for small areas in Wales. It identifies areas with the highest concentrations of several different types of deprivation. WIMD ranks all small areas in Wales from 1 (most deprived) to 1,909 (least deprived). It is a National Statistic produced by statisticians at the Welsh Government. Small areas are Census geographies called Lower-layer Super Output Areas (LSOAs).

Analysis of PPM prevalence LSOA and tenure type. *Tenure type* is categorised by housing that is: rented social and rented private housing and owner-occupied housing. Analysis of PPM prevalence MSOA by *emergency hospital admissions for Chronic Obstructive Pulmonary Disease (COPD)* (England 2021) and *emergency hospital admissions for all respiratory disease* in in Wales. COPD is the name used to describe a number of conditions including emphysema and chronic bronchitis. Deaths from COPD are a major global public health problem, particularly in countries and regions with higher levels of socioeconomic deprivation.[31]

Housing quality is measured by three variables: (1) proportion of properties with energy performance certificate rating D and above; (2) proportion of properties built 2007 and later; and (3) proportion of properties with multi-glazed windows per LSOA.

### Patient and public involvement

Patients and the public were not involved in the development of research questions, design of the study, recruitment, and conduct of the study, or dissemination of the study results.

## Results

Our dataset shows that England and Wales have on average 11% households using PPM, with higher frequency in Wales (14%) than England (11%). Regions with high PPM are concentrated in major cities such as London, Birmingham, Manchester as well as Northeast, the Wash, Cornwall, and Wales (especially North Wales and Cardiff) (Fig.1).

**Figure 1.**
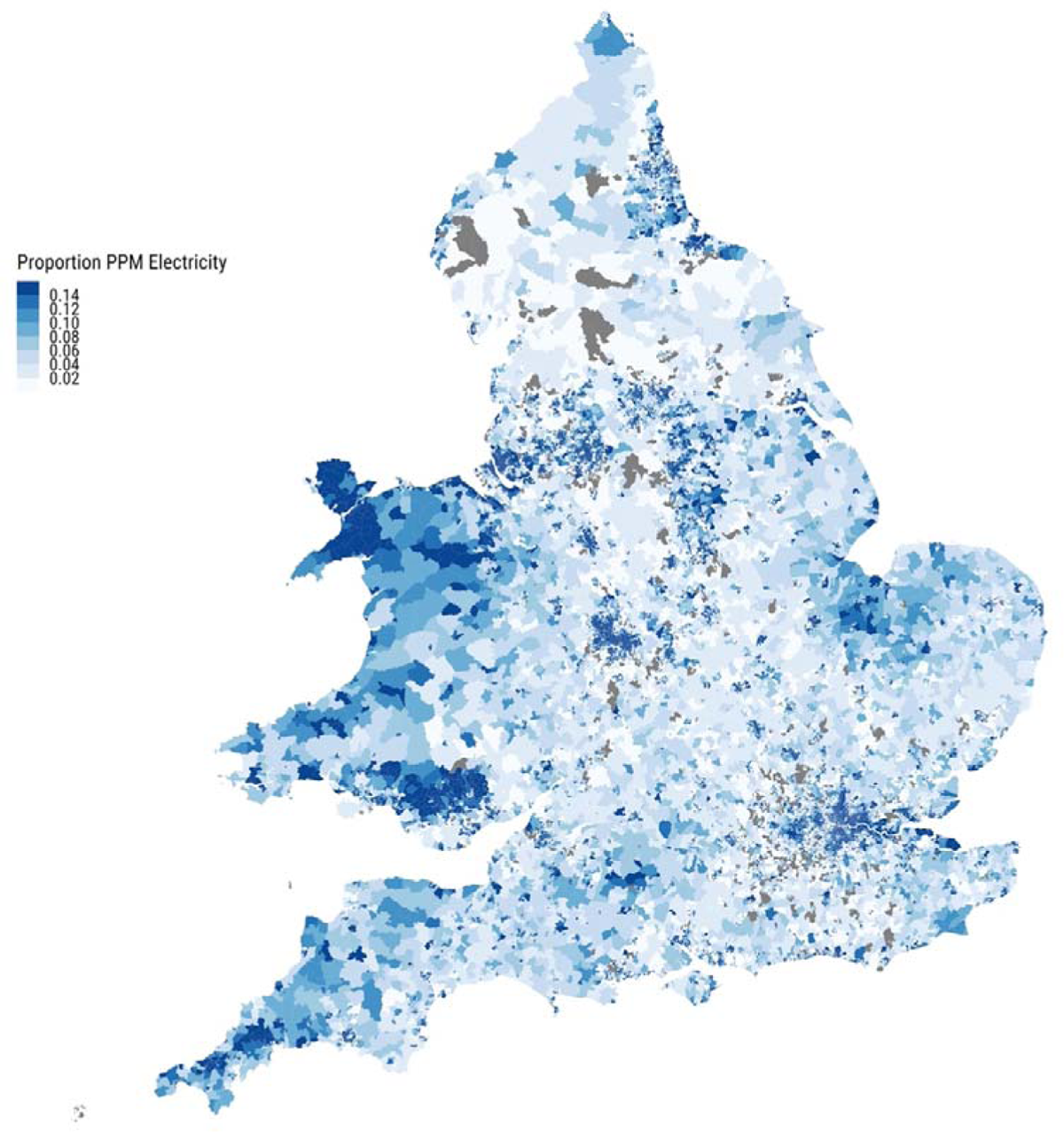
Sub-national (LSOA) percentages of prepayment electricity meters. England & Wales.

We then explored the associations between the PPM prevalence and multiple deprivation indices (Fig. 2, details are available on online supplemental table S1). PPM prevalence is strongly correlated (0.62 [0.62 to 0.63]) with the fuel poverty index in England, measured by the Low Income Low Energy Efficiency (LILEE) indicator. PPM prevalence was more strongly associated with deprivation indicators in every single domain compared to fuel poverty except for the living environment deprivation domain, which measures the quality of the indoor and outdoor environment. This is because fuel poverty takes into account house quality, measured by energy performance rating, and the living environment deprivation domain also measures housing quality. Higher PPM prevalence is strongly associated with having a: lower income (−0.83 [−0.83 to −0.82]), being in receipt of employment benefits (−0.76 [−0.77 to −0.76]), lower education (−0.69 [−0.70 to −0.69]) and higher levels of health deprivation (−0.65 [−0.65 to −0.64]). Higher PPM prevalence is also highly associated higher income deprivation affecting children (−0.8 [−0.81 to −0.80]) and for older people (−0.79 [−0.80 to −0.79]), which measures the actual proportion of children and older people living in income deprived families. A weak correlation was found between PPM and Barriers to Housing and services deprivation (−0.17 [−0.19 to −0.16]). The finding is somewhat surprising since one would expect PPM to be associated with physical and financial accessibility of housing. We therefore conducted a robustness analyses using housing quality data to disentangle the association between PPM and house quality in the next section.

**Figure 2.**
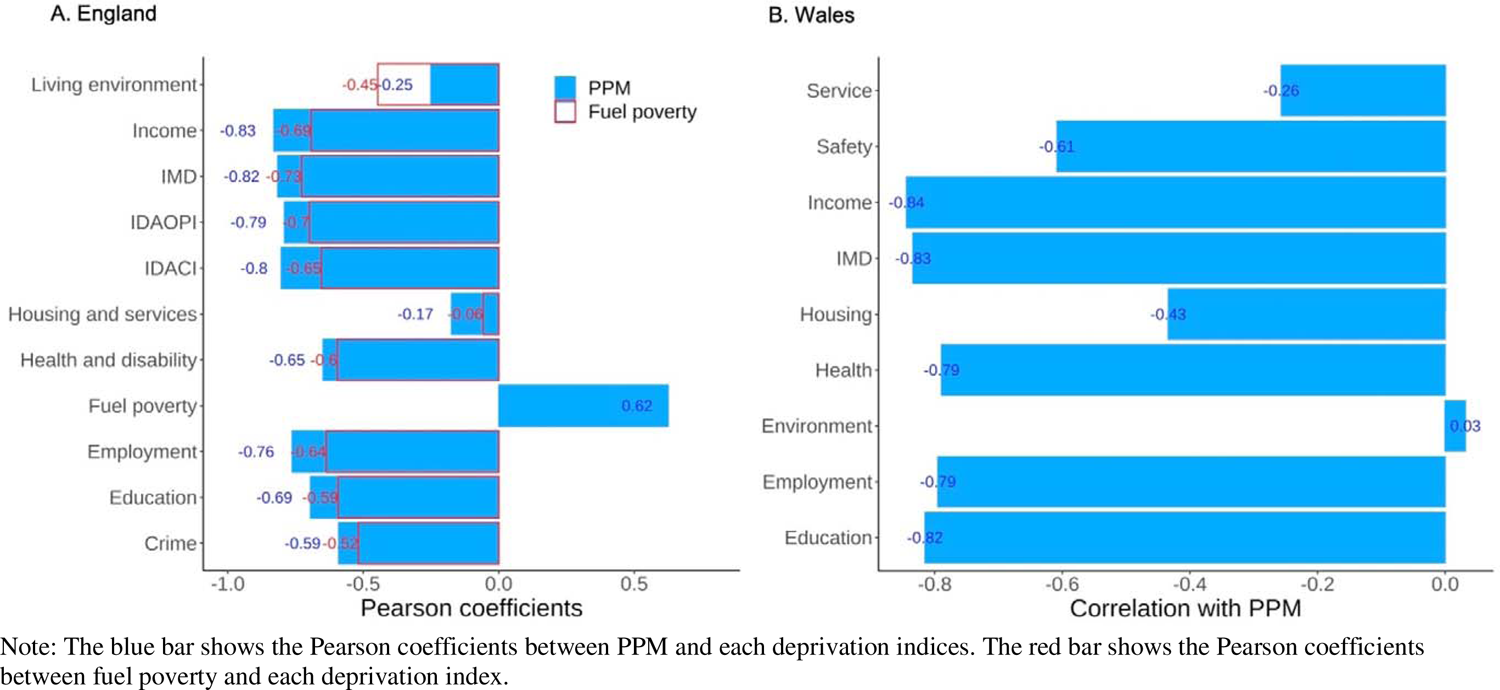
Correlation between prepayment electricity meters prevalence and multiple deprivation indices. England (A) and Wales (B). Note: The blue bar shows the Pearson coefficients between PPM and each deprivation indices. The red bar shows the Pearson coefficients between fuel poverty and each deprivation index.

Wales demonstrated the same pattern as England. The only exception was for the housing and the environment domains. The differences are due to how the two domains are measured in England and Wales. The housing domain in Wales accounts for housing quality, which is not measured in the Housing domain in England (as mentioned above, in England housing quality is measured in the living environment domain). Therefore, the association in housing and PPM was much stronger in Wales (−0.43 [−0.47 to −0.40]) than in England (−0.17 [−0.19 to −0.16]). Physical environment domain in Wales consists of three weighted domains of: air quality, flood risk and green space, which has little association with PPM (0.03 [−0.1 to 0.08]).

PPM prevalence is strongly associated with social rent (0.67 [0.66 to 0.68]) (Fig.3), suggesting that areas with higher PPM are more likely to be in social housing, which points to more targeted governmental interventions in this housing stock.

**Figure 3.**
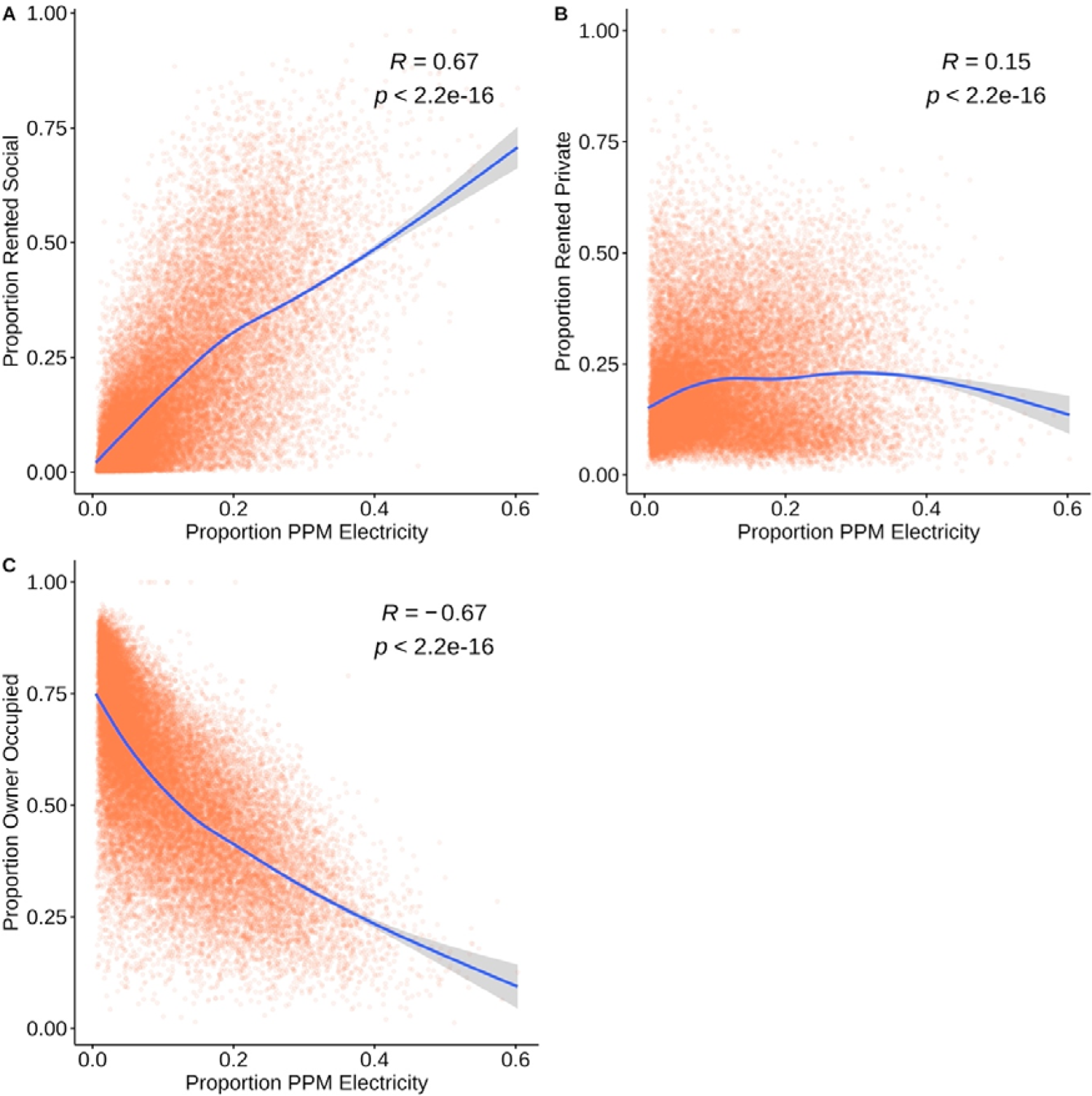
Prepayment electricity meters prevalence (LSOA) and tenure type. (A) Social rent. (B) Private rent. (C) Owner occupied.

Since a lack of heat has been shown to have extreme health consequences, we analysed the association between PPM prevalence and emergency hospital admission for respiratory diseases in England and Wales (Fig. 4). A higher proportion of PPMs is strongly associated with hospital admission for respiratory diseases in both England (0.69 [0.68 to 0.70]) and Wales (0.64 [0.58 to 0.70]), indicating that people living in PPM households are exposed to an even higher risk of illness. Given that these figures are prior to the energy crisis, our estimates are very high but still conservative in light of the upcoming winter and cost of living crisis.

**Figure 4.**
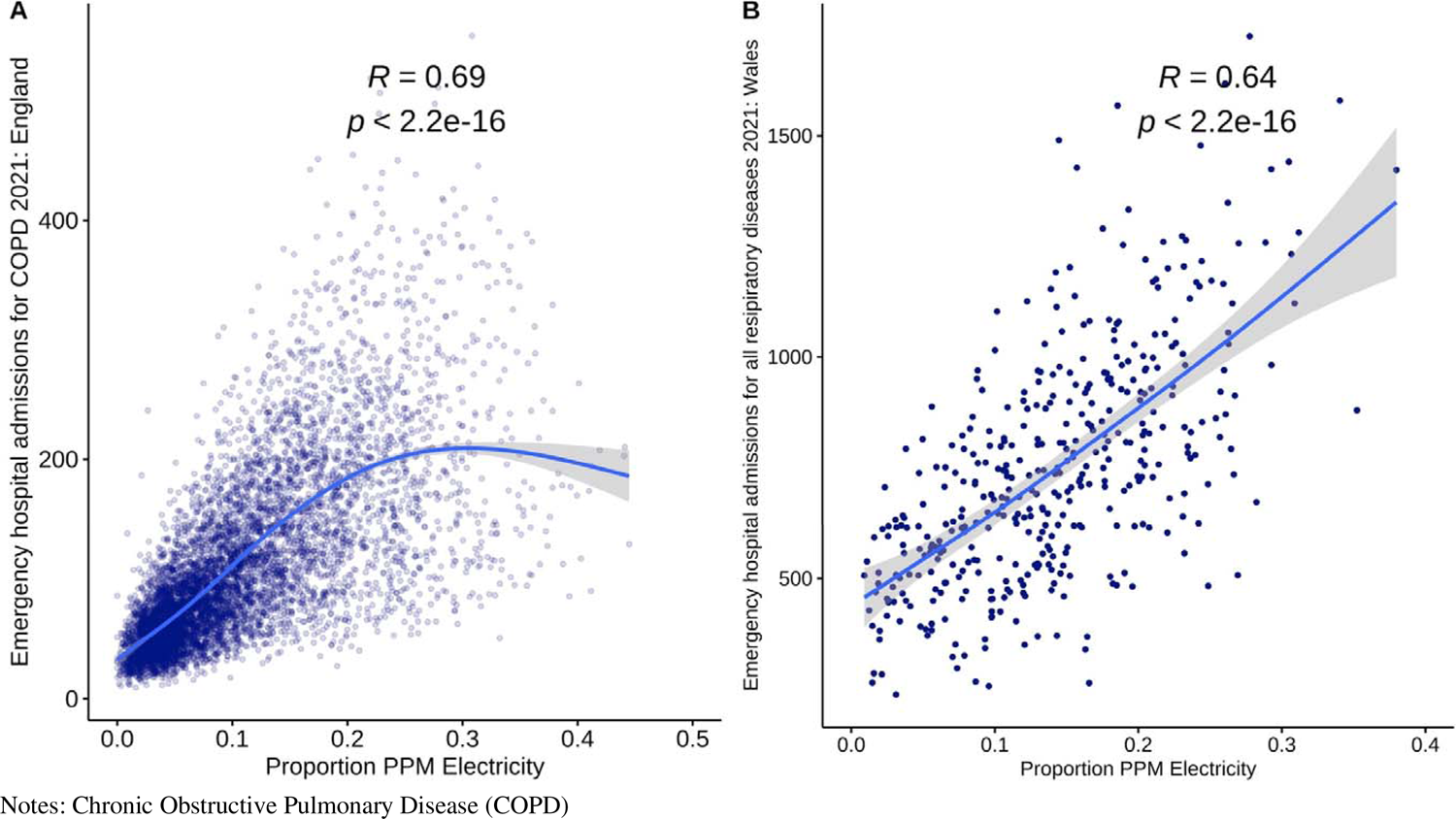
Prepayment electricity meters prevalence (MSOA) and emergency hospital admissions for COPD in England 2021 (A) and emergency hospital admissions for all respiratory diseases in Wales 2021 (B). Notes: Chronic Obstructive Pulmonary Disease (COPD)

To further examine the association between PPM and house quality, we examine the correlation between PPM and percentage Energy Efficiency Rating higher than D, property age, and multi-glazed windows per LSOA. We found no strong associations between PPM and housing quality variables (online supplementary table S1).

## Discussion

This study is the first to move beyond urgent editorial and calls to action to provide empirical evidence to demonstrate that PPMs are associated with fuel poverty and a variety of structural economic, social and health deprivation indicators. We found that PPMs (excluding smart meters with prepayment mode) are highly concentrated amongst those who already occupy the most vulnerable positions in society. This includes those with lower income and education, in receipt of employment benefits, and higher levels of health deprivation. A striking finding was that PPM prevalence is associated with households that have a higher proportion of children and older people living in income-deprived families. PPMs are also strongly associated with social housing (0.67 [0.66 to 0.68]), suggesting that the government has a responsibility to take an active role in this areas. Instead of blanket policies, such as the recent £400 per household Energy Bills Support Scheme that costly and lack sufficient targets,[32] we demonstrate that focussing on those who have PPMs can be an effective and efficient manner to reduce fuel poverty among a specific and easily identifiable group, who require urgent support to survive the winter. Our work also draws into question whether it is just and even counterproductive to force those who are already in debt or a vulnerable situation to a more expensive PPM system.

Households with PPMs can serve as a targeted and immediate intervention to cope with the energy crisis, cold weather and hospital admission emergency that is pushing a large group of vulnerable households into a ‘heat or eat’ dilemma. Although our study is informative for England and Wales, care should be taken when generalising our findings to other areas. We call for the release of PPM data in Scotland and Northern Ireland, and to include payment methods in social surveys to allow researchers and governments to evaluate risk for more focused prevention and preparedness.

## Data Availability

All data produced in the present study are available upon reasonable request to the authors

## Acknowledgements

MCM and all researchers are supported by an ERC Advanced Grant CHRONO (957566); the Leverhulme Trust Large Centre Grant for the Leverhulme Centre for Demographic Science and the ESRC/UKRI Connecting Generations Centre (ES/Vo14188/1). KD is supported by the Economic and Social Research Council grant ES/P000649/1.

## Conflict of Interests

All authors have completed the ICMJE uniform disclosure form at http://www.icmje.org/disclosure-of-interest/ and declare research funding support as above; MCM has received research grants from the European Research Council (ERC), ESRC/UKRI, Leverhulme Trust, Dutch Science Foundation (NWO); MCM has served on SAGE, is a Special Advisor to the European Commissioner of the Economy; a member of the No 10 Data Science Advisory Group and a member on the Scientific and Ethics Board of Our Future Health and Founder of Data2ThePeople; no other relationships or activities that could appear to have influenced the submitted work.

## Notes

### Competing Interest Statement

The authors have declared no competing interest.

### Summary of Updates

The methods section is updated to clarify the sample; Figure 4 revised; Supplemental files updated.

