## Supplementary material for "Prepayment meters strongly associated with economic and health deprivation: an observational, cross-sectional study": online supplemental

Fig S1. Distribution of IMD for LSOA with missing prepayment meters (PPM) in (A) England and (B) Wales.

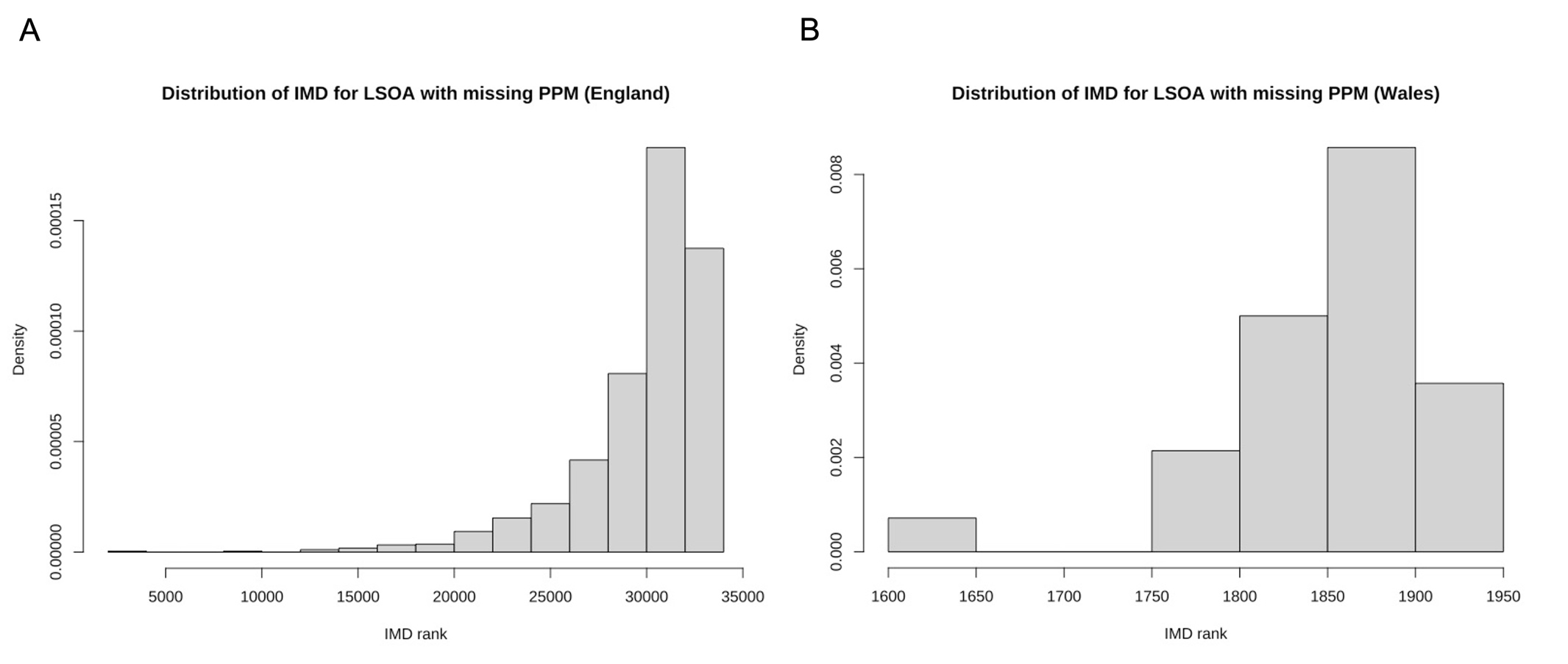

Table S1. Pearson correlation coefficients of prepayment meters (PPM) and social and health deprivation variables with 95% confidence interval.

|  | | N | PPM | 95% Confidence Interval |
| --- | --- | --- | --- | --- |
|  | England | | | |
| Fuel poverty | | 31,451 | 0.624 | (0.618, 0.631) |
| IMD | | 31,451 | -0.815 | (-0.819, -0.812) |
| Income | | 31,451 | -0.830 | (-0.833, -0.826) |
| Employment | | 31,451 | -0.762 | (-0.766, -0.757) |
| Education | | 31,451 | -0.695 | (-0.700, -0.689) |
| Health and Disability | | 31,451 | -0.648 | (-0.654, -0.641) |
| Crime | | 31,451 | -0.590 | (-0.598, -0.583) |
| Housing and services | | 31,451 | -0.174 | (-0.185, -0.164) |
| Living environment | | 31,451 | -0.251 | (-0.261, -0.240) |
| IDACI | | 31,451 | -0.802 | (-0.806, -0.798) |
| IDAOPI | | 31,451 | -0.791 | (-0.795, -0.786) |
| Emergency hospital admission for COPD^*^ | | 8,481 | 0.688 | (0.675, 0.700) |
|  | Wales (N=1,881) | | | |
| IMD | | 1,881 | -0.834 | (-0.847, -0.820) |
| Income | | 1,881 | -0.844 | (-0.857, -0.831) |
| Employment | | 1,881 | -0.795 | (-0.811, -0.778) |
| Health | | 1,881 | -0.789 | (-0.806, -0.772) |
| Education | | 1,881 | -0.815 | (-0.830, -0.799) |
| Service | | 1,881 | -0.257 | (-0.299. -0.214) |
| Housing | | 1,881 | -0.434 | (-0.470, -0.397) |
| Safety | | 1,881 | -0.608 | (-0.636, -0.579) |
| Environment | | 1,881 | 0.032 | (-0.014, 0.077) |
| Emergency hospital admission for all respiratory diseases^*^ | | 410 | 0.641 | (0.580,0.695) |
|  | England & Wales | | | |
| Social rent | | 30,891 | 0.669 | (0.663 0.675) |
| Private rent | | 31,462 | 0.148 | (0.138, 0.159) |
| Owner occupied | | 31,467 | -0.674 | (-0.680, -0.668) |
| Energy performance certificate D and above | | 31,477 | 0.235 | (0.225, 0.246) |
| Age of property (built after 2007) | | 27,002 | 0.022 | (0.010, 0.034) |
| Multi-glazing windows | | 31,471 | 0.093 | (0.082, 0.104) |

^*^ Middle Layer Super Output Area (MSOA) level analyses. Other outcomes are Lower Layer Super Output Area (LSOA) level analyses.

Figure S2. Data cleaning procedure.

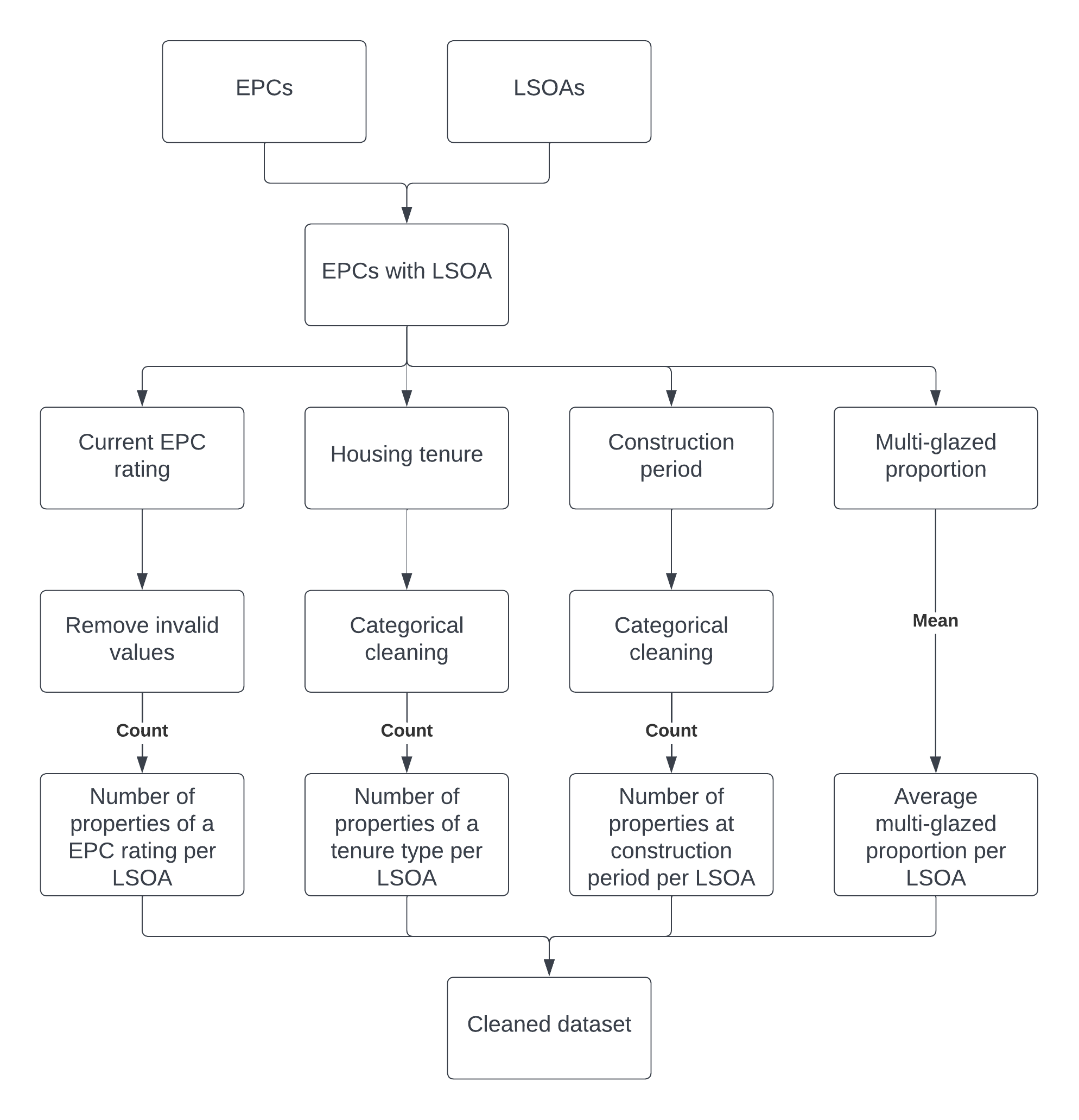
